# Scalable Risk Stratification for Heart Failure Using Artificial Intelligence applied to 12-lead Electrocardiographic Images: A Multinational Study

**DOI:** 10.1101/2024.04.02.24305232

**Authors:** Lovedeep S Dhingra, Arya Aminorroaya, Veer Sangha, Aline Pedroso Camargos, Folkert W Asselbergs, Luisa CC Brant, Sandhi M Barreto, Antonio Luiz P Ribeiro, Harlan M Krumholz, Evangelos K Oikonomou, Rohan Khera

## Abstract

**Background:** Current risk stratification strategies for heart failure (HF) risk require either specific blood-based biomarkers or comprehensive clinical evaluation. In this study, we evaluated the use of artificial intelligence (AI) applied to images of electrocardiograms (ECGs) to predict HF risk.

**Methods:** Across multinational longitudinal cohorts in the integrated Yale New Haven Health System (YNHHS) and in population-based UK Biobank (UKB) and Brazilian Longitudinal Study of Adult Health (ELSA-Brasil), we identified individuals without HF at baseline. Incident HF was defined based on the first occurrence of an HF hospitalization. We evaluated an AI-ECG model that defines the cross-sectional probability of left ventricular dysfunction from a single image of a 12-lead ECG and its association with incident HF. We accounted for the competing risk of death using the Fine-Gray subdistribution model and evaluated the discrimination using Harrel’s c-statistic. The pooled cohort equations to prevent HF (PCP-HF) were used as a comparator for estimating incident HF risk.

**Results:** Among 231,285 individuals at YNHHS, 4472 had a primary HF hospitalization over 4.5 years (IQR 2.5-6.6) of follow-up. In UKB and ELSA-Brasil, among 42,741 and 13,454 people, 46 and 31 developed HF over a follow-up of 3.1 (2.1-4.5) and 4.2 (3.7-4.5) years, respectively. A positive AI-ECG screen portended a 4-fold higher risk of incident HF among YNHHS patients (age-, sex-adjusted HR [aHR] 3.88 [95% CI, 3.63-4.14]). In UKB and ELSA-Brasil, a positive-screen ECG portended 13- and 24-fold higher hazard of incident HF, respectively (aHR: UKBB, 12.85 [6.87-24.02]; ELSA-Brasil, 23.50 [11.09-49.81]). The association was consistent after accounting for comorbidities and the competing risk of death. Higher model output probabilities were progressively associated with a higher risk for HF. The model’s discrimination for incident HF was 0.718 in YNHHS, 0.769 in UKB, and 0.810 in ELSA-Brasil. Across cohorts, incorporating model probability with PCP-HF yielded a significant improvement in discrimination over PCP-HF alone.

**Conclusions:** An AI model applied to images of 12-lead ECGs can identify those at elevated risk of HF across multinational cohorts. As a digital biomarker of HF risk that requires just an ECG image, this AI-ECG approach can enable scalable and efficient screening for HF risk.

## BACKGROUND

Despite the rising global burden of heart failure (HF) and the availability of evidence-based therapies for preventing and slowing the progression of the disease, there is a lack of a reliable approach for identifying individuals at the highest risk for developing HF.^1,2^ Due to this absence of an established and accessible screening strategy, patients often suffer the consequences of delayed diagnosis, including clinical HF, frequent hospitalizations, and premature mortality.^3–5^ Identifying individuals most likely to develop future HF can alleviate these risks with early initiation of low-cost medical therapies that have been proven in clinical practice guidelines to modify the trajectory of the disease, reducing both the risk for incident clinical HF and improving life expectancy.^6–9^

Several serum assay- and clinical score-based strategies have been proposed to predict incident HF.^10–18^ While serum assay-based biomarkers such as N-terminal pro–B-type natriuretic peptide (NT-proBNP) and high-sensitivity cardiac troponin I (hs-Tn) are independently associated with an elevated risk of incident HF,^19,20^ they are limited by the need for an invasive blood draw and frequent inaccessibility at the point-of-care.^15,18^ Predictive models based on clinical risk scores often require specialized testing and have varying predictive discrimination and feasibility of deployment.^11–13,21^ Recently, artificial intelligence (AI)-enhanced interpretation of electrocardiograms (ECGs; AI-ECG) has been proposed to detect hidden cardiovascular disease signatures from 12-lead ECGs.^22–30^ However, these deep learning models have focused on the cross-sectional detection of prevalent systolic dysfunction or HF,^27–34^ with limited application in predicting incident HF.^27,29,35,36^ Moreover, most current approaches use raw ECG voltage data as inputs, inaccessible to clinicians and patients at the point-of-care.^29,30^ Thus, there is an unmet need for practical and non-invasive screening tools that rely on ubiquitous and interoperable data sources to predict the future risk of HF.^27^ In our previous work, we reported an image-based AI-ECG screening approach with which a positive screen portended a higher risk of developing left ventricular systolic dysfunction (left ventricular ejection fraction [LVEF] < 40%) in patients with normal LVEF.^27^ However, an AI-ECG approach for comprehensive screening of HF risk is essential to realize the goals of HF prevention.

In this study, across three geographically and clinically distinct cohorts, we evaluated the hypothesis that an AI-ECG model developed to detect signatures of LV dysfunction on an ECG image at baseline will identify those at an elevated risk of new-onset HF.

## METHODS

The Yale Institutional Review Board approved the study protocol and waived the need for informed consent as the study involves secondary analysis of pre-existing data. An online version of the model is publicly available for research use at https://www.cards-lab.org/ecgvision-lv.

### Data Sources

We used data from the YNHHS, the UK Biobank (UKB) cohort, and the Brazilian Longitudinal Study of Adult Health (ELSA-Brasil), in our study (**Figure 1**). While the YNHHS represents a large and diverse healthcare system in the US, the UKB and ELSA-Brasil represent the largest population-based cohorts in the UK and Brazil, respectively, with protocolized baseline evaluation and detailed healthcare data capture. A brief overview of the data sources is included in the **Supplementary Methods**.

**Figure 1:**
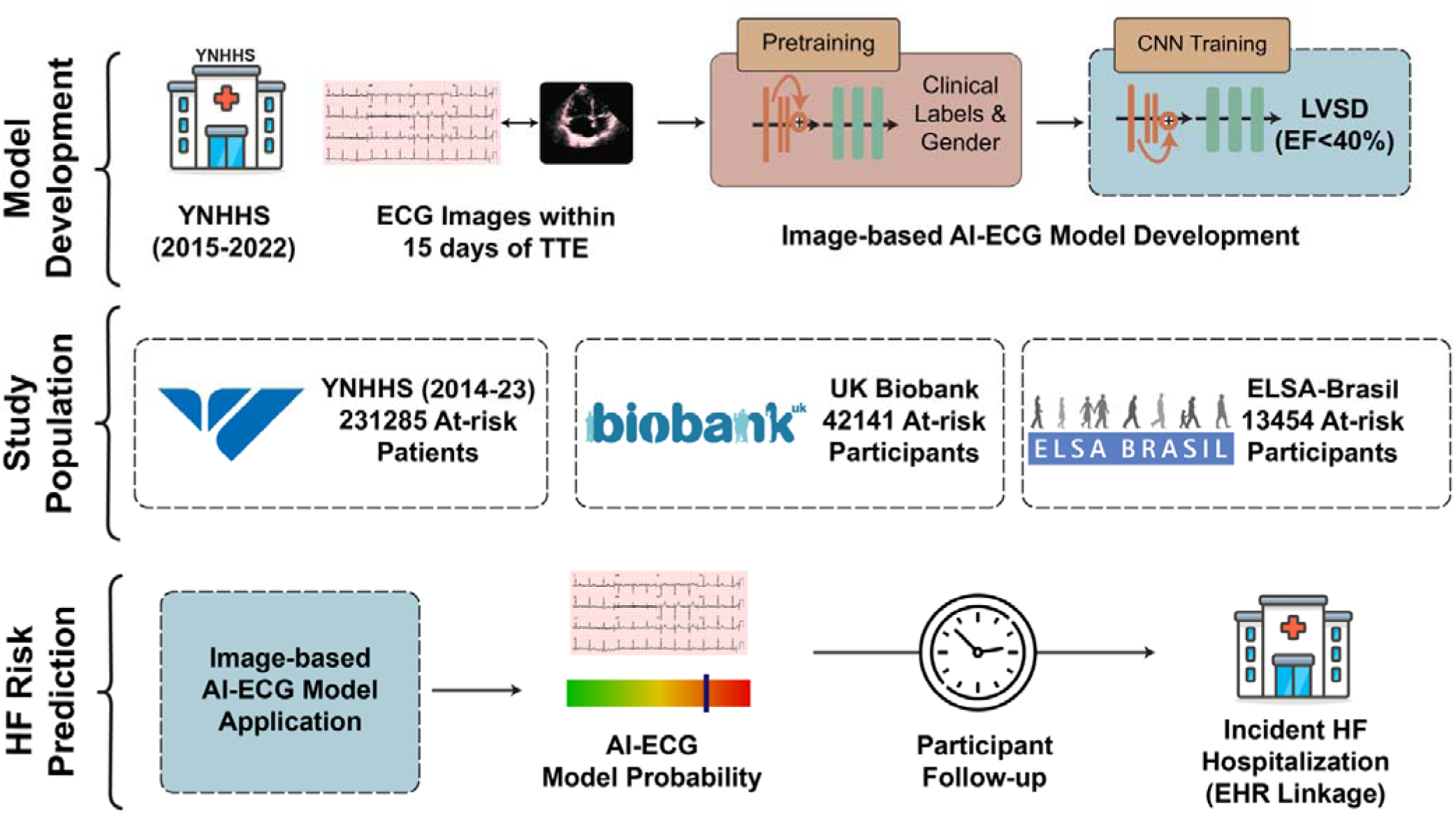
Study Overview. Abbreviations: AI, Artificial Intelligence; ECG, Electrocardiogram; EHR, Electronic Health Records; LVSD, Left Ventricular Systolic Dysfunction; UKBB; UK Biobank; YNHHS, Yale New Haven Health System

**Figure 2:**
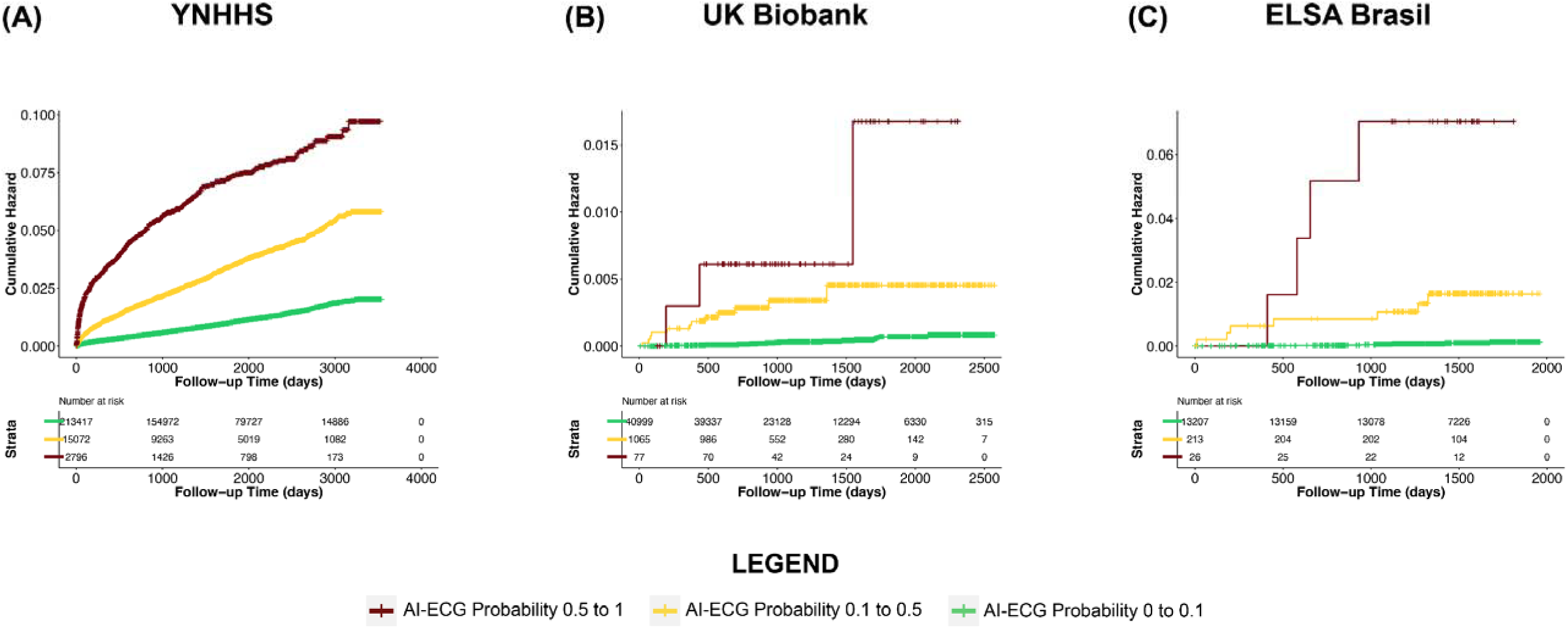
Age- and Sex-adjusted Cumulative Hazard Curves for Incident Heart Failure across Bins of Model Output Probability at (A) Yale New Haven Health System (B) UK Biobank. Abbreviations: AI-ECG, Artificial Intelligence-enhanced Electrocardiogram; SD, standard deviation

### Study Population

Across data sources, we included individuals with no known HF who had undergone a 12-lead ECG and followed them for the development of incident HF. For this, we constructed a cohort of patients seeking care at YNHHS, representing a large integrated EHR-based cohort along with well-characterized population-based cohort studies of UKB and ELSA-Brasil.

Among YNHHS patients, we identified the first recorded encounter for all patients within the EHR and instituted a 1-year blanking period to define prevalent HF (**Supplementary Methods; Figure S1**). Among 325,319 patients who had one or more ECGs after this 1-year blanking period, 76,736 patients who had been included in the AI-ECG model development, and 15,754 patients with any HF diagnosis code before their ECG were excluded. Moreover, 1544 patients with an echocardiogram with an LVEF below 50% or moderate or severe left ventricular diastolic dysfunction were also excluded (**Figure S2**).

In the UKB, we identified all 42,366 participants who had undergone a protocolized ECG as a part of the study procedures. Using the linked national EHR data from the UK, we excluded 225 participants who had been hospitalized with the record indicating an HF diagnosis code before the baseline ECG. Similarly, in ELSA-Brasil, we identified all 15,105 participants with a protocolized ECG at baseline, excluding 227 with HF at baseline and 58 with an LVEF of less than 50% noted on the baseline echocardiogram **(Figure S2)**.

### Study Exposure: AI-ECG-based HF Risk

The AI-ECG-based risk of HF represented the direct deployment of a previously developed AI-ECG model that detects left ventricular systolic dysfunction (defined as LVEF<40%) on ECG images,^27^ without any further development or fine-tuning.

Briefly, the model was developed in the YNHHS using ECG images from patients with paired echocardiograms and validated across 6 demographically diverse and geographically distinct populations as a cross-sectional association between AI-ECG-based and imaging-based evaluation of left ventricular systolic dysfunction (area under the receiver operating characteristic curve of 0.91 [95% CI, 0.90–0.92]). Further information about the application of the model in this study is included in the **Supplementary Methods**. The study exposure was a positive AI-ECG screen, defined as a model output probability greater than 0.1, representing the threshold at which the model achieved a sensitivity of over 90% for the cross-sectional detection of systolic dysfunction during internal validation.^27^

### Study Outcomes and Covariates

The study outcome was new-onset or incident HF. In the YNHHS, this was defined as an inpatient admission with an International Classification of Disease Tenth Revision Code – Clinical Modification (ICD-10-CM) for HF as the principal hospitalization diagnosis (**Table S1**). The choice of this approach was guided by the over 95% specificity of HF diagnosis codes, especially as the principal discharge diagnosis, for a clinical diagnosis of HF.^37^ We pursued the same approach in UKB, where we used linked National Health Service EHR to identify hospitalization records with HF as the principal diagnosis code. In ELSA-Brasil, incident HF was identified either by in-person interview or the annual telephonic surveillance for all hospitalizations, followed by independent medical record review and adjudication of HF hospitalizations by two cardiologists (**Supplementary Methods**).^38^

To evaluate the specificity of the HF risk defined by the AI-ECG model, we examined the risk of other cardiovascular conditions, including acute myocardial infarction (AMI), stroke hospitalizations, and all-cause mortality (**Table S1**).

Information about all-cause death was defined by established approaches for each source (**Supplementary Methods**). A composite outcome of major adverse cardiovascular events (MACE) was defined as any primary HF, AMI, or stroke hospitalization, or death.

For all analyses, common demographic covariates were selected across cohorts, including age, sex, race, and ethnicity. Age was defined at the time of the index ECG across all cohorts. We further identified the presence of hypertension and type 2 diabetes mellitus using encounters for these conditions in the YNHHS EHR as well as the EHR records linked with UKB (**Table S1**). In ELSA-Brasil, information about demographic covariates and baseline hypertension and type 2 diabetes was recorded at the baseline study visit.^39^ Race, or skin color, was self-classified based on Brazil’s National Bureau of Statistics definition and classified as White, Black, “Pardo”, Asian, or Others.^39,40^

### Study Comparator

Across all study cohorts, we compared the predictive performance of the AI-ECG model with the pooled cohort equations to prevent HF (PCP-HF), representing sex- and race-specific clinical risk models for estimating incident HF risk, developed and validated using data from 7 population-based cohorts.^11^ The PCP-HF risk score includes a combination of several demographic and laboratory-based covariates, including age, body mass index, systolic blood pressure, total cholesterol, high-density cholesterol, fasting blood glucose, smoking status, antihypertensive medication use, antihyperglycemic medication use, as well as electrocardiographically defined QRS duration. The PCP-HF input features were defined across the study cohorts using the EHR and study visits (**Supplementary Methods**).^41–44^

### Statistical Analysis

Categorical variables were reported as counts and percentages, and continuous variables as median and interquartile range (IQR). The association of AI-ECG-based risk with incident HF was evaluated in age- and sex-adjusted Cox proportional hazard models with time-to-first HF event as the dependent variable and the AI-ECG-based screen status (positive or negative) as the key independent variable. Further, to account for the competing risk of death while evaluating incident HF, we used age- and sex-adjusted multi-outcome Fine-Gray subdistribution hazard models.^45^ The discrimination of AI-ECG and PCP-HF for HF prediction was assessed using Harrel’s c-statistic, which incorporates the time dependence of outcomes and the non-linearity in the association between predictions and time-to-outcomes.^46–48^ The statistical significance level was set at 0.05. All statistical analyses were executed using Python 3.11.2 and R version 4.2.0.

## RESULTS

A total of 231,285 individuals constituted the study cohort at YNHHS, with a median age of 57 years (IQR 42, 70) and 130,941 (56.6%) were women, and 85,559 (37.0%) were non-White. Over a median follow-up of 4.5 years (IQR 2.5-6.6), 4472 (1.9%) had a primary HF hospitalization, 9645 (4.2%) had a primary HF hospitalization or an echocardiogram with a LVEF under 50% on subsequent echocardiogram, and 17,542 (7.5%) died **(Table 1)**.

**Table 1.** Population Characteristics of the Study Cohorts. Abbreviations: AMI, acute myocardial infarction; ECG, Electrocardiogram; ELSA-Brasil, Brazilian Longitudinal Study of Adult Health; HF, heart failure; IQR, Interquartile Range; UKB, UK Biobank; YNHHS, Yale New Haven Health System

| Characteristic |  | YNHHS | UKB | ELSA-Brasil |
| --- | --- | --- | --- | --- |
| Number |  | 231285 | 42141 | 13454 |
| Age at ECG, Median [IQR] |  | 57.4 [42.1,70.2] | 65 [59,71] | 51 [45,58] |
| Female Sex, N (%) |  | 130941 (56.6) | 21795 (51.7) | 7348 (54.6) |
| Race/Ethnicity, N (%) | White | 145726 (63.0) | 40691 (96.6) | 6920 (51.4) |
|  | Black | 36605 (15.8) | 304 (0.7) | 2130 (15.8) |
|  | Hispanic | 36298 (15.7) | - | - |
|  | Asian | 4221 (1.8) | 600 (1.4) | 332 (2.5) |
|  | Other | 2565 (1.1) | 546 (1.3) | 305 (2.3) |
|  | Brazilian “Pardo” | - | - | 3767 (28.0) |
|  | Missing | 5870 (2.5) | - | - |
| Death, N (%) |  | 17380 (7.5) | 346 (0.8) | 229 (1.7) |
| Follow-up Time, Years; Median [IQR] |  | 4.5 [2.5,6.6] | 3.1 [2.1,4.5] | 4.2 [3.7, 4.5] |
| Positive Screens, N (%) |  | 17868 (7.7) | 1142 (2.7) | 239 (1.8) |
| Primary HF hospitalization during follow-up, N (%) |  | 4472 (1.9) | 46 (0.1) | 31 (0.2) |
| Primary HF hospitalization or an echocardiogram with LVEF < 50% during follow-up, N (%) |  | 9645 (4.2) | - | - |
| Any HF hospitalization during follow-up, N (%) |  | 19004 (8.2) | 231 (0.5) | - |
| Any HF hospitalization or an echocardiogram with LVEF < 50% during follow-up, N (%) |  | 21849 (9.4) | - | - |
| Primary AMI hospitalization during follow-up, N (%) |  | 288 (0.1) | 208 (0.5) | 60 (0.4) |
| Primary Stroke hospitalization during follow-up, N (%) |  | 3688 (1.6) | 210 (0.5) | 59 (0.4) |
| Major Adverse Cardiovascular Events during follow-up, N (%) |  | 24059 (10.4) | 768 (1.8) | 338 (2.5) |

In UKB, 42,141 included participants had a median age of 65 years (IQR 59-71), 21795 (51.7%) were women, and 40,691 (96.6%) were of White race. Over a median follow-up of 3.1 years (IQR 2.1-4.5), 46 (0.1%) had a HF hospitalization event and 346 (0.8%) died **(Table 1)**.

In ELSA-Brasil, the median age of the 13,454 included participants was 51 years (IQR 45-58) and 7348 (54.6%) women. A total of 31 people developed HF, and 229 died over a median follow-up of 4.2 years (IQR 3.7-4.5).

### Predicting the Risk of Incident HF

At YNHHS, 17,868 (7.7%) patients screened positive based on the AI model applied to ECG images. A positive screen was associated with over 6.5-fold higher risk of incident HF (HR 6.51 [95% CI, 6.11-6.93]; **Table 2)**. Patients with a positive AI-ECG screen had a nearly 4-fold risk of incident HF, compared with patients with a negative screen, after accounting for differences in age and sex (adjusted HR [aHR], 3.88 [95% CI, 3.63-4.14]), as well as additionally accounting for differences baseline HF risk factors of hypertension and diabetes (aHR 3.73 [95% CI, 3.50-3.99]). Accounting for the competing risk of death, in addition to age and sex, a positive screen was associated with an aHR of 3.54 (95% CI, 3.30-3.79) for incident HF **(Table 2)**.

**Table 2.** Model Performance for Predicting Heart Failure Risk. Abbreviations: ELSA-Brasil, Brazilian Longitudinal Study of Adult Health; HTN, hypertension; T2DM, type-2 diabetes mellitus; UKB, UK Biobank; YNHHS, Yale New Haven Health System

| Model | Covariates | YNHHS | UKB | ELSA-Brasil |
| --- | --- | --- | --- | --- |
| **Cox Proportional Hazard Model** | Positive Screen | 6.51 (6.11-6.93) | 18.33 (9.90-33.97) | 32.06 (15.36-66.92) |
| **Cox Proportional Hazard Model** | Positive Screen + Age + Sex | 3.88 (3.63-4.14) | 12.85 (6.87-24.02) | 23.50 (11.09-49.81) |
| **Cox Proportional Hazard Model** | Positive Screen + Age + Sex + HTN + T2DM | 3.73 (3.50-3.99) | 11.36 (6.04-21.36) | 17.36 (8.55-35.26) |
| **Fine-Gray Subdistribution Hazard Model** | Positive Screen + Age + Sex + Competing Risk of Death | 3.54 (3.30-3.79) | 12.70 (6.70-24.07) | 22.79 (10.21-50.89) |
| **Fine-Gray Subdistribution Hazard Model** | Positive Screen + Age + Sex + HTN + T2DM + Competing Risk of Death | 3.41 (3.18-3.65) | 11.14 (5.72-21.70) | 17.96 (8.14-39.61) |

The association of a positive screen with an elevated risk of HF was noted across demographic subgroups of age, sex, race, and ethnicity **(Table S2)**. Notably, a positive screen portended an 8-fold higher risk of incident HF in patients < 65 years of age at the time of ECG (aHR 8.00 [95% CI, 7.12-8.99]).

The model performance was consistent in sensitivity analyses with subsets of the population with (1) ≥3 years of follow-up in the EHR (aHR, 3.75 [95% CI, 3.46-4.08]), and (2) ≥1 encounter every 2 years (aHR, 3.49 [95% CI, 3.25-3.75]). Further, the patterns were consistent when a random ECG was chosen instead of the first ECG (aHR, 3.76 [95% CI, 3.38-4.18]) and across different definitions of HF **(Tables S3 and S4)**.

In UKB, 1142 (2.7%) participants screened positive with the AI-ECG model. A positive AI-ECG screen portended an 18-fold higher hazard for incident HF (HR 18.33 [95% CI, 9.90-33.97]). Accounting for age, sex, baseline hypertension, and type 2 diabetes, screen-positive participants had an 11-fold higher risk of HF (HR 11.36 [95% CI, 6.04-21.36]; **Table 2)**. Further, this risk was even higher in individuals below 65 years of age, with an age- and sex-adjusted HR of 25.63 (95% CI, 6.34-103.61; **Table S2)**. After accounting for the competing risk of death, a positive screen was associated with a nearly 13-fold risk of incident HF (aHR: 12.70; 95% CI, 6.70-24.07).

At ELSA-Brasil, 239 (1.8%) participants had a positive AI-ECG screen, with a 24-fold higher risk for incident HF (age- and sex-adjusted HR 23.50 [95% CI, 11.09-49.81]) compared with screen-negative participants. This association was consistent even after accounting for the competing risk of death (aHR of 22.79; 95% CI, 10.21-50.89; **Table 2**).

### Hazard Across Model Probability Outputs

In the YNHHS cohort, each 0.1 increment in the model output probability portended a 36% higher hazard of an incident primary HF hospitalization (aHR 1.36 [95% CI, 1.35-1.38]). Among screen-positive patients, patients with model output probability between 0.1-0.5 and 0.5-1 had over 3- and 7-fold higher risk of incident HF, compared with screen-negative patients (aHR 3.31 [95% CI, 3.08-3.55] and 7.11 [95% CI, 6.42-7.88], respectively). Higher model probabilities were progressively associated with a higher risk of incident HF across various probability bins **(Table S5).**

These patterns were replicated across both UKB and ELSA-Brasil, with a 0.1 increase in model probability associated with 81% and 93% higher risk of incident HF (aHR 1.81 [95% CI, 1.58-2.07] and aHR 1.93, [95% CI, 1.68-2.21], respectively, **Table S5)**. A higher threshold for defining a screen-positive ECG consistently defined a higher hazard of incident HF (**Table S6**).

### Prediction of other cardiovascular outcomes

A positive AI-ECG screen was also associated with a more modest but still elevated risk of AMI (aHR, 1.44; 95% CI, 1.04-2.00), all-cause death (aHR 1.19 [95% CI, 1.15-1.24]), or MACE, defined by AMI, stroke, HF, or death (aHR 2.10 [95% CI, 2.04-2.17]; **Table S4**) with the YNHHS. Similarly, in the UKB, a positive AI-ECG screen was associated with an elevated risk for AMI and stroke (aHRs 3.16 [95% CI, 1.98-5.02] and 2.30 [95% CI, 1.36-3.9], respectively), all-cause death (aHR 2.13 [95% CI, [1.41-3.24]), and MACE (aHR 2.79 [95% CI, 2.17-3.6]; **Table S4)**. This pattern was replicated in ELSA-Brasil across all non-HF cardiovascular outcomes of AMI (aHR 3.53 [95% CI, 1.4-8.85]), stroke (aHR 5.74 [95% CI, 2.59-12.72]), death (aHR 3.64 [95% CI, 2.27-5.83]), and MACE (aHR 4.04 [95% CI, 2.77-5.89]), with a comparably smaller increase in risk of non-HF cardiovascular events than HF risk.

### Comparison with PCP-HF

In YNHHS, the AI-ECG model had a model discrimination based on Harrel’s c-statistic of 0.718 (0.697-0.738), compared with 0.601 (0.581-0.621) for PCP-HF score (p <0.001; **Table 3)**. In UKB and ELSA-Brasil, the AI-ECG’s model discrimination for incident HF was 0.769 (95% CI, 0.670-0.867) and 0.810 (95% CI, 0.714-0.907), respectively, comparable to that for PCP-HF (UKB: p = 0.71; ELSA-Brasil: p = 0.89). Across all cohorts, incorporating model probability with PCP-HF yielded a statistically significant improvement in discrimination over the use of PCP-HF alone (YNHHS: 0.147 [95% CI, 0.124-0.170]; UKB: 0.127 [95% CI, 0.032-0.223]; ELSA-Brasil: 0.106 [95% CI, 0.030-0.181]; **Table 3**).

**Table 3.** Comparison of Discrimination for AI-ECG Model Output Probability and Pooled Cohort equations to Prevent Heart Failure for Incident Heart Failure. Abbreviations: ELSA-Brasil, Brazilian Longitudinal Study of Adult Health; PCP-HF, Pooled Cohort equations to Prevent Heart Failure; UKB, UK Biobank; YNHHS, Yale New Haven Health System.

| Covariates | YNHHS |  |  | UKB |  |  | ELSA-Brasil |  |  |
| --- | --- | --- | --- | --- | --- | --- | --- | --- | --- |
|  | Harrel's c-statistic | Marginal difference over Harrel's c-statistic for PCP-HF | P-value | Harrel's c-statistic | Marginal difference over Harrel's c-statistic for PCP-HF | P-value | Harrel's c-statistic | Marginal difference over Harrel's c-statistic for PCP-HF | P-value |
| PCP-HF | 0.601<br>(0.581-0.621) | - | - | 0.740<br>(0.647-0.834) | - | - | 0.821<br>(0.748-0.893) | - | - |
| AI-ECG Model Output Probability | 0.718<br>(0.697-0.738) | 0.117<br>(0.086-0.147) | <0.001 | 0.769<br>(0.670-0.867) | 0.028<br>(-0.119-0.175) | 0.71 | 0.810<br>(0.714-0.907) | 0.010<br>(-0.149-0.130) | 0.89 |
| AI-ECG Model Output Probability + Age + Sex | 0.724<br>(0.705-0.743) | 0.122<br>(0.098-0.148) | <0.001 | 0.832<br>(0.756-0.910) | 0.092<br>(-0.014-0.198) | 0.09 | 0.881<br>(0.820-0.942) | 0.060<br>(0.039-0.159) | 0.23 |
| AI-ECG Model Output Probability + PCP-HF | 0.748<br>(0.730-0.766) | 0.147<br>(0.124-0.170) | <0.001 | 0.868<br>(0.810-0.926) | 0.127<br>(0.032-0.223) | 0.008 | 0.926<br>(0.886-0.966) | 0.106<br>(0.030-0.181) | 0.006 |

## DISCUSSION

Across three clinically and geographically distinct cohorts, a deep learning model that was developed to define signatures of left ventricular systolic dysfunction on ECG images represents a non-invasive and accessible digital biomarker for predicting HF risk from ECG images as the sole input. A positive AI-ECG screen was associated with a 4- to 24-fold higher hazard of incident HF across different populations than a negative screen, consistent after accounting for competing risk of death. The model was evaluated in demographically and racially diverse cohorts, with high predictive performance across subgroups of age, sex, race, and ethnicity. We observed a progressive increase in risk based on the LV dysfunction probability on baseline ECG, such that graded increments in the model output probability were associated with higher hazards of incident HF. A positive screen was also relatively specific for HF risk, with a 2-to-6-fold higher risk of HF than the risk of other cardiovascular outcomes, such as MACE and death. In addition to being substantially easier to use than PCP-HF that uses an ECG-derived measure in addition to substantial laboratory and clinical testing, the AI-ECG approach had incremental discrimination over PCP-HF. Therefore, deep learning-enhanced interpretation of ECG images can represent a scalable and reliable strategy for risk stratification for incident HF.

The ability to identify individuals at the highest risk for HF is crucial given the high clinical and economic impact of HF, along with the availability of evidence-based risk-mitigating therapies.^6–9^ Thus, there has been substantial investigation into defining risk for HF.^12–18^ Notably, blood-based biomarkers have been a key focus on investigation, with an NT-proBNP of 125pg/ml associated with 2.4-fold higher risk of incident HF.^10,18–20^ This is further potentiated with the addition of hs-Tn.^18,49^ However, the application of these serum-based assays for HF risk is limited by the need for blood draws and laboratory testing.^15,50^ Similarly, PCP-HF, and other proposed HF risk scores, such as the Atherosclerosis Risk in Communities HF score or the Heart ABC model require detailed clinical history, physical examination, and testing for routine and specialized laboratory measures,^8,11,16,21,51^ such that these are not consistently recommended in clinical practice guidelines.

While neural network-based models have previously been designed to detect prevalent systolic dysfunction or HF,^27–34^ our study suggests the role of an AI-ECG model as a biomarker for new-onset HF. This AI-based approach can enable opportunistic HF screening for patients undergoing clinical ECGs and also facilitate population-based screening approaches for HF.^5^ Moreover, given that ECGs are most commonly available to clinicians and patients as digital images or printouts, the application of conventional signal-based AI-ECG models has been limited by access to raw input ECG voltage data.^29–34^ The image-based approach can use interoperable digital images or smartphone photographs of printouts, representing a scalable strategy for deployment in the community.^27^

The predictive performance of our model was high across demographic subgroups, with the highest predictive power in younger individuals across cohorts. This suggests an opportunity for proactive HF screening in younger individuals, followed by implementing risk-mitigating strategies. Further, progressive increases in the AI-ECG score were associated with a progressively higher risk of HF. This dose-dependence represents ideal characteristics for a predictive biomarker, enabling graded risk stratification and proactive mitigation in those at the highest predicted risk.^14^

Our study has certain limitations that merit consideration. First, outcomes in the EHR are prone to inconsistent capture due to site-specific variability in coding practices,^52,53^ though we opted for a specific definition of HF. The association of a positive AI-ECG screen with the risk of HF across YNHHS and population-based cohorts at UKB and ELSA-Brasil indicates that the model captures a predictive signature of disease across a spectrum of disease phenotypes. Moreover, the patterns were consistent across several sensitivity analyses that defined these conditions differently in YNHHS and UKB. Further, in ELSA-Brasil, HF events were expert-adjudicated using established clinical criteria. Second, despite an integrated health system with broad geographic coverage, some HF and other outcome events may have occurred outside YNHHS, which may be reflected in the lower risk of HF and other events compared with the UKB and ELSA-Brasil, where the follow up was consistent and protocolized. Therefore, the risk of HF observed for patients in the YNHHS likely represents the lower bound of their actual HF risk, based on the findings in the prospective cohorts. Moreover, the patients who underwent ECG testing were selected, with a risk for the unmeasured potential risk profile of those who underwent a clinical ECG but had a negative AI-ECG screen. Finally, while the study finds a high risk of subsequent HF, it is unclear whether the risk of HF identified by AI-ECG is modifiable. Nevertheless, these observations may suggest a focus on targeted identification and management of known HF risk factors.

## CONCLUSION

An AI model applied to images of 12-lead ECGs can identify those at elevated risk of HF across multinational cohorts. As a digital biomarker of HF risk that requires just an ECG image, this AI-ECG approach can enable scalable and efficient screening for HF risk.

## Data Availability

Data from the Yale New Haven Health System represent protected health information that cannot be shared publicly. Data from the UK Biobank and the ELSA-Brasil are available to licensed users.

