## Supplementary material for "Scalable Risk Stratification for Heart Failure Using Artificial Intelligence applied to 12-lead Electrocardiographic Images: A Multinational Study": Online Supplement

**SUPPLEMENTARY MATERIALS**

**SUPPLEMENTARY METHODS**

**Data Sources**

The YNHHS is the largest referral center in southern New England, comprising a network of 5 hospitals and a large outpatient medical group, serving areas of Connecticut, New York, and Rhode Island. The electronic health record (EHR) data represent an extract from the Clarity database, comprising the corpus of healthcare information acquired during patient care at YNHHS using Epic.^52,54^ Data between 2014 through 2023 was included in the study.

We used data from the UKB under research application #71033. The UKB is a prospective observational study of 502,468 people aged 40-69 years recruited in 2006-2010.^55^ These participants underwent ECG recordings starting 2014 and were followed through till 2021. The UKB represents the largest population-based cohort in the United Kingdom with protocolized imaging as a part of the study and EHR linkage from the National Health Service.^41,56^

The ELSA-Brasil study is a large multicenter prospective cohort from Brazil, enrolling 15,105 volunteering participants aged 35-74 years at recruitment between the years 2008-2010.^39,57^ These participants represent active and retired civil servants from six higher education and research institutions in Brazilian state capitals in three geographical regions of the country: Southeast (Belo Horizonte, Rio de Janeiro, São Paulo and Vitória), South (Porto Alegre) and Northeast (Salvador).^58^ ELSA’s main objectives are to investigate the development and progression of chronic diseases and their determinants in the Brazilian adult population. Baseline data were collected at the time of recruitment using interviews with previously validated questionnaires and clinical, laboratory and imaging exams including protocolized ECGs and echocardiograms.^39,57^ In-person follow-up visits were conducted every three to four years to ascertain exposure status and to identify changes in baseline subclinical and clinical parameters. In addition, all participants were interviewed yearly via telephone to obtain information on new diagnoses, hospitalization, and death with adjudicated clinical events based on expert medical record review.^57^

**Study Population – Cohort Identification at YNHHS**

To identify patients with prevalent HF at the time of ECG, we identified the first recorded encounter for all patients within the EHR and followed for 1 year. Patients with prevalent HF based on either a diagnosis code for HF or an echocardiogram with reduced LVEF (defined as LVEF<50%) or left ventricular diastolic dysfunction (defined as “moderate” or “severe” left ventricular diastolic dysfunction) we excluded from the study. The baseline ECG for patients was defined after this 1-year blanking period to exclude prevalent HF (**Figure S1**). The YNHHS cohort also excluded patients previously included in the development of the AI-ECG algorithm and the small proportion of individuals who opted out of research participation (<0.01% of all YNHHS patients).

We further constructed subpopulations in the YNHHS cohort with more consistent follow-up to ensure the completeness of outcome assessments. This included patients with (1) ≥3 years of follow-up in YNHHS and (2) ≥1 encounter within the healthcare system every 2 years. As a sensitivity analysis, we also identified a single random ECG (instead of the first ECG) for each patient after the 1-year blanking period following cohort entry.

**Study Exposure: AI-ECG-based HF Risk**

The image-based AI-ECG model had 89% sensitivity and 77% specificity for identifying patients with concomitant left ventricular systolic dysfunction on cardiac imaging.^27^

In this study, the model was deployed on ECG images with a voltage calibration of 10 mm/mV in a standard layout, with the limbs and precordial leads arranged in four columns of 2.5-second each, representing leads I, II, and III; aVR, aVL, and aVF; V1, V2, and V3; and V4, V5, and V6. A 10-second recording of the lead I signal was included as a rhythm strip. These images were converted to greyscale and down-sampled to 300x300 pixels using Python Image Library.^59^

While the study exposure was a positive-screen ECG (model output probability > 0.1), we further defined graded thresholds based on AI-ECG probabilities of 0-0.1, 0.1-0.3, 0.3-0.5, 0.5-0.7, and 0.7-1 to evaluate the association of a higher risk score with incident HF.

**Study Outcomes and Covariates**

In ELSA-Brasil, incident HF was identified either by in-person interview or the annual telephonic surveillance and investigated by a designated committee that contacted health providers and requested copies of medical records for all hospitalizations. After investigation, the cardiovascular events were adjudicated by an independent review of two cardiologists. A third senior cardiologist defined the event in case of disagreement.^60^ Incident HF was identified from hospitalization records, based on the presence of a clinical diagnosis of HF, with the individual receiving pharmacological therapy for HF, in addition to any of the following: (1) pulmonary congestion on chest X-ray, (2) reduced ejection fraction or systolic dysfunction observed on cardiac imaging, or (3) preserved ejection fraction with evidence of moderate to severe diastolic dysfunction.

Information about all-cause death was available in the YNHHS EHR, with in-hospital mortality data supplemented from the Connecticut death index to improve capture of out-of-hospital patient mortality. Similarly, information about mortality was available in the UKB via linkage to the EHR and the UK national death registries. Information about death in the ELSA-Brasil study was recorded via telephonic surveillance and confirmed using the national mortality database and death certificates.

Among patients in the YNHHS, we also identified echocardiograms with an LVEF less than 50% performed after the ECG recording. Further, to evaluate the AI-ECG model for a different definition for incident HF, we also identified hospitalizations with any HF diagnosis in YNHHS and UKB.

**Study Comparator**

The PCP-HF represent sex- and race-specific equations for estimating 10-year risk of incident HF and include a variety of demographic and clinical features such as age, body mass index, systolic blood pressure, total cholesterol, high-density cholesterol, fasting blood glucose, current smoking status, antihypertensive medication use, antihyperglycemic medication use, and electrocardiogram measurement of QRS duration. To align with the score development, across cohorts, the PCP-HF score was calculated for White and Black individuals between 30 and 80 years of age with complete documentation of the score covariates. The calculated 10-year risk score was adjusted based on the length of follow-up for each individual to estimate the risk of heart failure over the study period.

In YNHHS, PCP-HF features were extracted from the EHR. Body mass index, systolic blood pressure, and laboratory measurements closest to and within two years of the ECG acquisition date were used for calculation. In the UKB, the demographic features were identified from the baseline visit. Blood pressure measurement and smoking status assessment were conducted at the time of ECG acquisition. Laboratory values were measured in the first and second study visits, while ECGs were recorded in third and fourth visits.^42^ We used the laboratory values closest to the ECG acquisition for the calculation of PCP-HF score. History of hypertension and diabetes were defined using ICD diagnosis codes from the linked EHR and self-reported use of anti-hypertensive and anti-hyperglycemic medications was recorded at the time of ECG acquisition.^41^ In ELSA-Brasil, all PCP-HF features, including the ECG recording, were captured at the baseline visit using established study protocols.^43,44^

**Figure S1. Overview of Cohort Creation at the Yale New Haven Health System.**

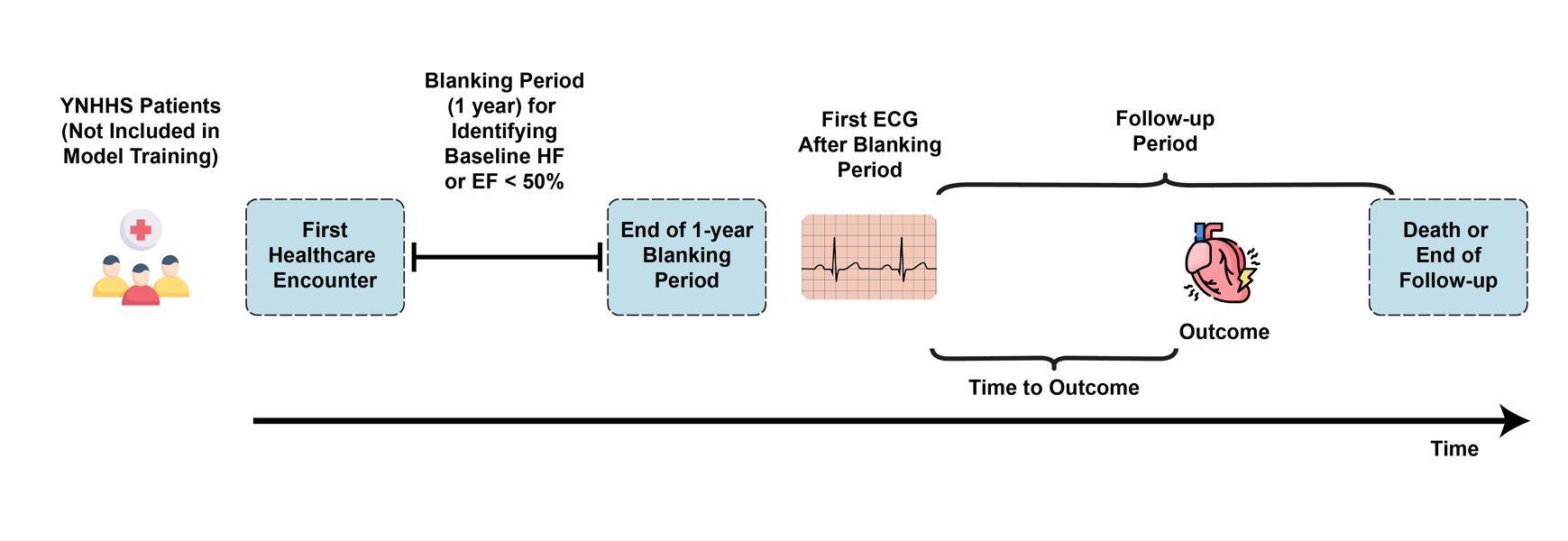

**Figure S2. Consort Diagram for (A) Yale New Haven Health System Cohort and (B) UK Biobank Cohort.** Abbreviations: AI, Artificial Intelligence; ELSA-Brasil, Brazilian Longitudinal Study of Adult Health; ECG, Electrocardiogram; HF, Heart Failure

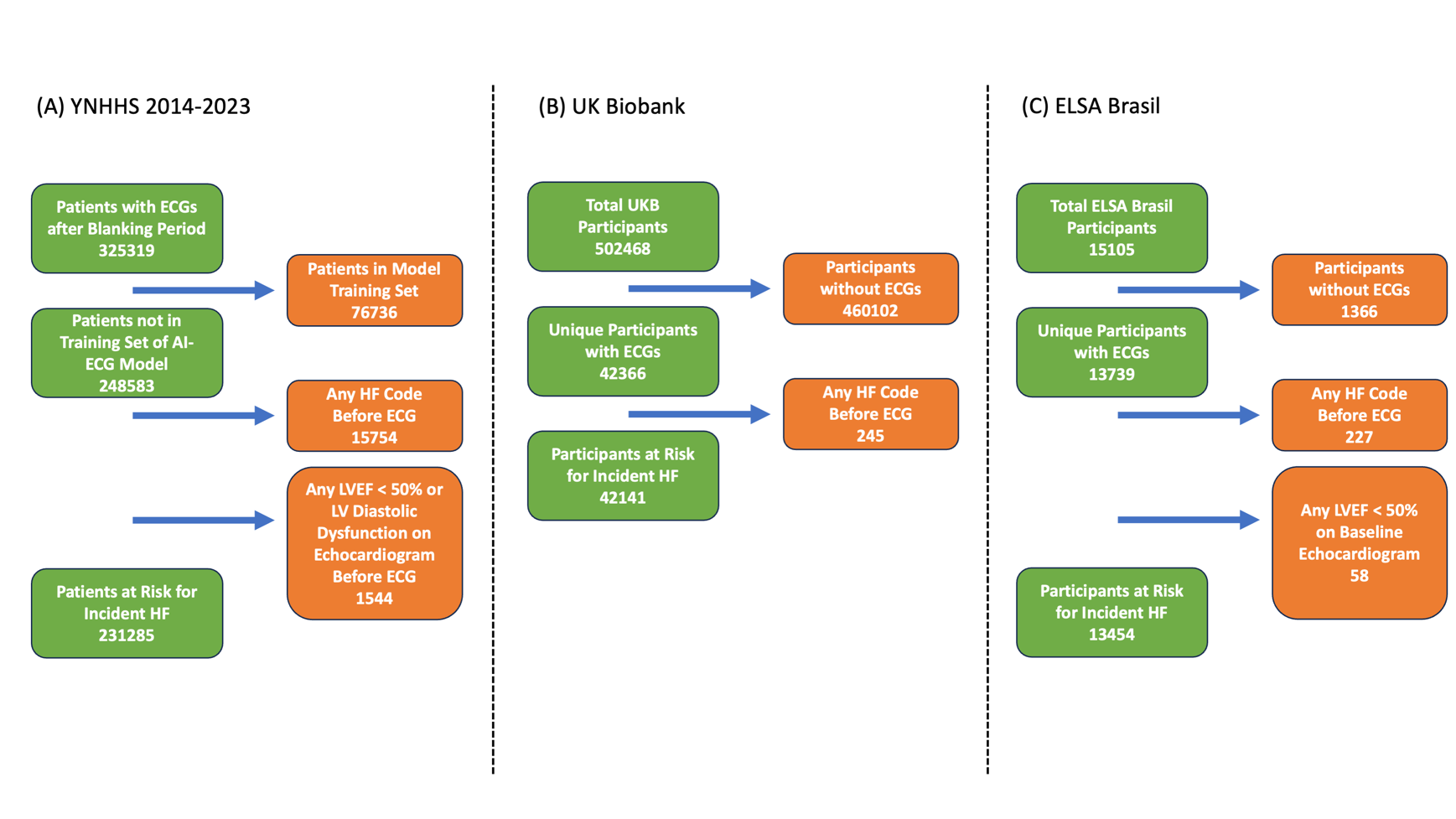

**Figure S3. Age- and Sex-adjusted Cumulative Hazard Curve for Primary Heart Failure Hospitalization within the Yale New Haven Health System Cohort, including a Random Electrocardiogram following the One-year Blanking Period.**

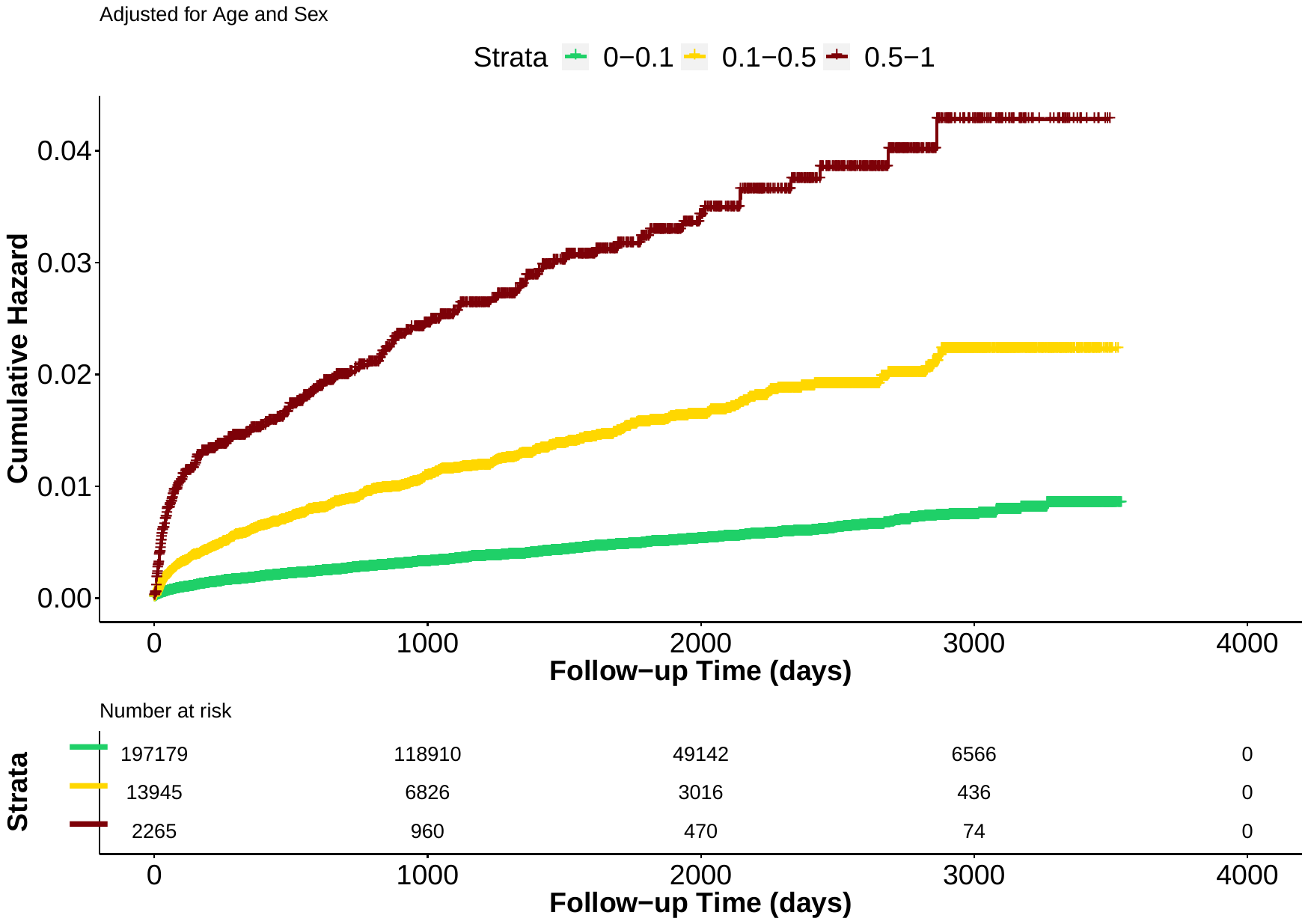

**Table S1. International Classification of Disease Tenth Revision Codes for the Identification of Comorbidities and Outcomes.** Abbreviations: ICD-10-CM, International Classification of Disease Tenth Revision Clinical Modification Codes.

| **Condition** | **ICD-10-CM codes** |
| --- | --- |
| **Heart Failure** | ‘I11.0’,’I13.0’,’I13.2’,’I50’,’I50.0’,’I50.1’,’I50.9’,’Z95.81’,’I09.81’ |
| **Acute Myocardial Infarction** | ‘I21’, ‘I22’, ‘I23’, ‘I24.0’, ‘I24.8’, ‘I24.9’ |
| **Stroke** | ‘G45’,’G45.0’,’G45.1’,’G45.2’,’G45.3’,’G45.4’,’G45.8’,’G45.9’,  ‘I63’,’I63.0’,’I63.1’,’I63.2’,’I63.3’,’I63.4’,’I63.5’,’I63.8’,’I63.9’,’I64’,  ‘I65’,’I65.0’,’I65.1’,’I65.2’,’I65.3’,’I65.8’,’I65.9’,’I66’,’I66.0’,’I66.1’,  ‘I66.2’,’I66.3’,’I66.4’,’I66.8’,’I66.9’,’I67.2’,’I69.3’,’I69.4’ |
| **Type 2 Diabetes Mellitus** | ‘E11’,’E11.0’,’E11.1’,’E11.2’,’E11.3’,’E11.4’,’E11.5’,’E11.6’,  ‘E11.7’,’E11.8’,’E11.9’,’O24.1’ |
| **Hypertension** | ‘I10’,’I11’,’I11.0’,’I11.9’,’I12’,’I12.0’,’I12.9’,  ‘I13’,’I13.0’,’I13.1’,’I13.2’,’I13.9’,’I67.4’,  ‘O10’,’O10.0’,’O10.1’,’O10.2’,’O10.3’,’O10.9’,’O11’ |

**Table S2. Model Performance for Prediction of Heart Failure Risk Across Demographic Subgroups.** Abbreviations: CI, Confidence Interval; ELSA-Brasil, Brazilian Longitudinal Study of Adult Health; HF, Heart Failure; UKB, UK Biobank; YNHHS, Yale New Haven Health System

| **Subgroup** | | **YNHHS** | | | **UKB** | | | **ELSA-Brasil** | | |
| --- | --- | --- | --- | --- | --- | --- | --- | --- | --- | --- |
|  |  | **Total Number of Individuals at Risk** | **Number of Incident HF Events** | **Age- and Sex- Adjusted Cox Proportional Hazard Ratios (95% CI)** | **Total Number of Individuals at Risk** | **Number of Incident HF Events** | **Age- and Sex- Adjusted Cox Proportional Hazard Ratios (95% CI)** | **Total Number of Individuals at Risk** | **Number of Incident HF Events** | **Age- and Sex- Adjusted Cox Proportional Hazard Ratios (95% CI)** |
| **Age < 65** | | 151199 (65.4) | 1352 | 8.00 (7.12-8.99) | 20802 (49.4) | 9 | 25.63 (6.34-103.61) | 12038 (89.4) | 21 | 25.35 (10.01-64.21) |
| **Age ≥ 65** | | 80086 (34.6) | 3120 | 3.00 (2.78-3.24) | 21345 (50.6) | 37 | 11.1 (5.54-22.23) | 1416 (10.6) | 10 | 19.94 (5.60-70.99) |
| **Female** | | 130941 (56.6) | 2225 | 3.41 (3.09-3.76) | 21795 (51.7) | 11 | 13.41 (3.53-50.96) | 7348 (54.6) | 11 | 16.30 (6.20-42.89) |
| **Male** | | 100341 (43.4) | 2247 | 4.34 (3.98-4.74) | 20346 (48.3) | 35 | 12.87 (6.33-26.15) | 6106 (45.4) | 20 | 43.74 (12.98-147.42) |
| **Race/Ethnicity** | **White** | 145726 (63.0) | 3343 | 3.38 (3.13-3.64) | 40691 (96.6) | 46 | 12.85 (6.87-24.02) | 6920 (51.4) | 15 | 27.18 (9.34-79.07) |
|  | **Black** | 36605 (15.8) | 624 | 5.43 (4.59-6.42) | 304 (0.7) | 0 | - | 2130 (15.8) | 9 | 26.18 (6.65-103.05) |
|  | **Hispanic** | 36298 (15.7) | 358 | 5.73 (4.51-7.29) | 0 | - | - | - | - | - |
|  | **Asian** | 4221 (1.8) | 47 | 9.63 (5.17-17.96) | 600 (1.4) | 0 | - | 332 (2.5) | 0 | - |
|  | **Other** | 2565 (1.1) | 35 | 4.41 (2.03-9.57) | 546 (1.3) | 0 | - | 305 (2.3) | 0 | - |
|  | **Brazilian “Pardo”** | - | - | - | - | - | - | 3767 (28.0) | 7 | 7.89 (0.94-66.19) |
|  | **Missing** | 5870 (2.5) | 65 | 5.44 (3.16-9.39) | 0 | - | - | - | - | - |

**Table S3. Model Performance for Prediction of Incident Heart Failure Across Key Subsets of Interest.** Abbreviations: YNHHS, Yale New Haven Health System

| **Characteristic** | | **YNHHS with minimum 3-year follow-up** | **YNHHS with at least one encounter every two years** | **YNHHS Random ECG after Blanking Period** |
| --- | --- | --- | --- | --- |
| **Total Number of Patients at Risk** | | 94848 | 128466 | 213389 |
| **Number of Patients Developing Incident HF** | | 2898 | 3749 | 1685 |
| **Cox Proportional Hazard Model** | Positive Screen + Age + Sex | 3.75 (3.46-4.08) | 3.49 (3.25-3.75) | 3.76 (3.38-4.18) |

**Table S4. Age- and Sex- Adjusted Cox Proportional Hazard Models for the Prediction of Clinical Outcomes.** Abbreviations: AMI, acute myocardial infarction; HF, heart failure; UKB, UK Biobank; YNHHS, Yale New Haven Health System

| **Outcome** | **Age- and Sex- Adjusted Cox Proportional Hazard Models** | | |
| --- | --- | --- | --- |
|  | **YNHHS**  **Hazard Ratio (95% CI)** | **UKB**  **Hazard Ratio (95% CI)** | **ELSA-Brasil**  **Hazard Ratio (95% CI)** |
| **Primary HF Hospitalization** | 3.88 (3.63-4.14) | 12.85 (6.87-24.02) | 23.50 (11.09-49.81) |
| **Primary HF Hospitalization or an Echocardiogram with LVEF < 50%** | 5.05 (4.83-5.27) | - | - |
| **Primary AMI Hospitalization** | 1.44 (1.04-2.00) | 3.16 (1.98-5.02) | 3.53 (1.4-8.85) |
| **Primary Stroke Hospitalization** | 1.05 (0.95-1.17) | 2.30 (1.36-3.9) | 5.74 (2.59-12.72) |
| **All-cause Death** | 1.19 (1.15-1.24) | 2.13 (1.41-3.24) | 3.64 (2.27-5.83) |
| **Major Adverse Cardiovascular Events** | 2.10 (2.04-2.17) | 2.79 (2.17-3.6) | 4.04 (2.77-5.89) |
| **Any Hospitalization with HF** | 3.11 (3.00-3.22) | 7.32 (5.3-10.09) | - |
| **Any Hospitalization with HF or an Echocardiogram with LVEF < 50%** | 3.48 (3.37-3.59) | - | - |

**Table S5. Age- and Sex- Adjusted Hazard Ratios for Incident Heart Failure across Model Output Probabilities.** Abbreviations: ELSA-Brasil, Brazilian Longitudinal Study of Adult Health; UKB, UK Biobank; YNHHS, Yale New Haven Health System

| **Age- and Sex- Adjusted Cox Proportional Hazard Models** | | **YNHHS**  **Hazard Ratio (95% CI)** | **UKB**  **Hazard Ratio (95% CI)** | **ELSA-Brasil**  **Hazard Ratio (95% CI)** |
| --- | --- | --- | --- | --- |
| **Per 0.1 increase in model output probability** | | 1.36 (1.35-1.38) | 1.81 (1.58-2.07) | 1.93 (1.68-2.21) |
| **Grouped by ranges of model output probabilities** | **0-0.1** | Reference | Reference | Reference |
|  | **0.1-0.5** | 3.29 (3.06-3.54) | 11.30 (5.75-22.18) | 16.80 (7.02-40.25) |
|  | **0.5-1** | 7.16 (6.44-7.95) | 29.17 (8.82-96.48) | 80.11 (26.96-237.97) |
| **Grouped by ranges of model output probabilities** | **0-0.1** | Reference | Reference | Reference |
|  | **0.1-0.3** | 3.00 (2.77-3.26) | 10.27 (4.86-21.71) | 18.66 (7.44-46.85) |
|  | **0.3-0.5** | 4.38 (3.87-4.95) | 16.29 (4.93-53.83) | 10.68 (1.41-80.79) |
|  | **0.5-0.7** | 6.09 (5.30-7.00) | 22.44 (5.32-94.62) | 31.24 (4.16-234.78) |
|  | **0.7-0.9** | 7.63 (6.46-9.00) | 74.59 (10.05-553.42) | 170.11 (48.09-601.77) |
|  | **0.9-1** | 17.58 (13.28-23.27) | - | - |

**Table S6. Age- and Sex-adjusted Cox Proportional Hazard Models Across Different Model Output Probability Threshold for Defining a Positive Screen.** Abbreviations: ELSA-Brasil, Brazilian Longitudinal Study of Adult Health; UKB, UK Biobank; YNHHS, Yale New Haven Health System.

| **Model Output Probability Threshold for Positive Screen** | **Age- and Sex- Adjusted Cox Proportional Hazard Models** | | |
| --- | --- | --- | --- |
|  | **YNHHS**  **Hazard Ratio (95% CI)** | **UKB**  **Hazard Ratio (95% CI)** | **ELSA-Brasil**  **Hazard Ratio (95% CI)** |
| **>0.05** | 3.25 (3.06-3.46) | 6.93 (3.83-12.53) | 11.71 (5.64-24.36) |
| **>0.1** | 3.88 (3.63-4.14) | 12.85 (6.87-24.02) | 23.50 (11.09-49.81) |
| **>0.2** | 4.32 (4.01-4.66) | 14.46 (6.67-31.34) | 22.68 (9.12-56.40) |
| **>0.5** | 5.57 (5.03-6.17) | 20.88 (6.42-67.95) | 58.16 (20.07-168.48) |
